# Demographic and Clinical Correlates of Discordant QuantiFERON TB Gold Tuberculosis Screening Results in a Low Endemicity Setting

**DOI:** 10.1101/2025.09.02.25334912

**Authors:** Gaurav K. Sharma, Farah Haq, Arthur H. Totten, Luis A. Marcos, Charles Kyriakos Vorkas

## Abstract

Interferon-ɣ Release Assays (IGRAs), such as the QuantiFERON-TB Gold Plus (QFTTB) and T-SPOT.TB, are commonly used to detect prior exposure to *Mycobacterium tuberculosis* complex (*Mtb*), the causative agent of tuberculosis (TB). IGRA positive (IGRA+) asymptomatic individuals are diagnosed with presumed latent tuberculosis infection (LTBI) and often offered therapy to prevent progression to active disease. However, discordant results during serial or confirmatory IGRA testing pose challenges for interpretation and may lead to unnecessary LTBI treatment. We conducted a retrospective study of subjects who received QFTTB testing at Stony Brook Medicine between October 2020 and March 2024, focusing on discordant serial testing, to identify sociodemographic and clinical variables associated with quantitative QFTTB results. Variables measured included age, sex, race, comorbidities, and medication use. A total of 743 subjects were analyzed, including all 436 QFTTB-positive (QFTTB+) cases of 11,641 tests ordered (3.7%), of whom 16 were diagnosed with active TB. A random sample of 307 age-sex-matched QFTTB-negative controls were included. Of 203 subjects undergoing serial QFTTB testing, 170 (83.7%) had concordant results, while 33 (16.3%) showed discordance—23 (69.7%) with reversion and 10 (30.3%) with conversion. Conversions occurred in significantly older subjects (mean age 51.1 ± 15.0 vs. 37.0 ± 15.6, p = 0.025) and over longer intervals (415.1 vs. 91.2 days, p = 0.026). Comorbidities including cardiovascular disease, infections, and diabetes correlated with changes in NIL, TB1, and TB2 values. These findings highlight inconsistencies in QFTTB testing that complicate LTBI management and underscore the importance of confirmatory testing in low-incidence settings.

**Importance Statement:** Reliable interpretation of interferon-γ release assays (IGRAs) is critical for the diagnosis and management of latent tuberculosis infection (LTBI). However, variability in test performance, particularly during serial or confirmatory testing, complicates clinical decision-making and may result in unnecessary treatment. Our study demonstrates that demographic factors, clinical comorbidities, and testing intervals contribute to discordant QuantiFERON-TB Gold Plus (QFTTB) results. These findings underscore the need to integrate epidemiologic risk, clinical history, and repeat testing before initiating therapy, especially in low-incidence regions where the pre-test probability of infection is low. Improved understanding of IGRA variability can strengthen both patient care and research applications, including TB vaccine and biomarker studies.

## Introduction

Tuberculosis (TB) is a significant public health challenge, with an estimated 10.8 million new cases and 1.25 million deaths annually worldwide^1^. A cornerstone of TB control efforts is early detection of *Mycobacterium tuberculosis* complex (*Mtb*) exposure in asymptomatic individuals using immunodiagnostic screening and treatment of presumed latent tuberculosis infection (LTBI). This intervention is thought to reduce the 10% lifetime risk of progression to active TB and mitigate potential transmission to naïve contacts^2^. Accurate screening for LTBI is essential for public health strategies aimed at TB prevention with goal of elimination of *Mtb* in human reservoirs.

Interferon-ɣ Release Assays (IGRAs), such as the QuantiFERON-TB Gold Plus (QFTTB) and T-SPOT.TB, have emerged as alternatives to the traditional Tuberculin Skin Test (TST) for diagnosis of LTBI, with guidelines favoring IGRA testing to TST for TB screening^3^. These blood-based assays measure the release of interferon-ɣ (IFN-γ), a cytokine secreted by T lymphocytes upon recognition of *Mtb* antigens, offering improved specificity over TST by minimizing cross-reactivity with Bacillus Calmette-Guérin (BCG) vaccination and most non-tuberculous mycobacteria^4^. QFTTB is an IGRA that utilizes two antigen tubes —TB1 and TB2 — each containing synthetic peptide pools derived from early secreted antigenic target-6 (ESAT-6) and culture filtrate protein-10 (CFP-10), which are absent from BCG strains and some non-tuberculous mycobacteria^4,5^. While the TB1 tube is optimized to stimulate CD4^+^ T-helper cells restricted by major histocompatibility complex class II (MHC II), responses may also be elicited from CD8⁺ T-cells restricted by MHC I^6^. The TB2 tube includes additional shorter peptide pools designed to enhance sensitivity by stimulation of both CD4⁺ and CD8⁺ T cells^6,7^. The assay includes a mitogen tube as a positive control and a NIL tube to measure basal IFN-γ levels. Interpretation is based on the change in IFN-γ concentration measured by enzyme-linked immunoassay (ELISA) of whole blood compared to NIL control, using pre-defined cutoff thresholds^8^. If the mitogen tube IFN-γ concentration does not reach the positive control threshold, the result is reported as “indeterminate” and the antigen tubes are not tested. This assay has been widely incorporated into TB screening programs in both high- and low-incidence settings and is used as a primary measure of efficacy in experimental preventive TB vaccine clinical trials^4,9,10^. For example, in one Phase III clinical trial, IGRA conversion was used to measure incident primary infection in otherwise baseline asymptomatic IGRA negative individuals in high-risk settings^10^.

Despite its utility, serial IGRA testing can also demonstrate discordant results that may not accurately measure *Mtb*-specific immune responses. This raises concerns about assay reproducibility and reliability as a screening test, particularly in regions of low endemicity ^11,12^. For example, a person may test negative and subsequently positive with no clear evidence of recent *Mtb* exposure^12–14^. In turn, confirmatory IGRA testing may yield a discordant negative result. This variability is particularly concerning for individuals undergoing serial TB screening, where discordant results may misclassify individuals, leading to missed opportunities to initiate LTBI therapy or inappropriate treatment^14–16^. Moreover, discordant QFTTB testing as a measure of efficacy in clinical trials may confound the development of new TB vaccines^13^. Multiple factors have been reported to contribute to discordant IGRA results, including a dynamic host immune response, assay cutoff thresholds, and specimen processing^17,18^. Importantly, 8-19% of microbiologically confirmed active TB cases have negative IGRA results, attributed to T cell exhaustion during chronic infection^17^. It has also been reported that individuals with recent viral infections, malnutrition, or chronic inflammatory diseases may experience periods of immunosuppression, potentially leading to false-negative results^17,19^. Assay-specific variability has also been documented, with studies showing that different IGRA platforms (QFTTB vs. T-SPOT.TB) may yield conflicting results in the same individual^20^. Our study examined the hypothesis that sociodemographic and clinical variables correlate with QFTTB results, focusing on discordant serial testing in a community-based population in a low-endemicity setting in Suffolk County, Long Island, NY that reports 3 cases/100,000 per year^21^.

## Methods

This was a retrospective electronic medical record (EMR) review of subjects who underwent routine standard of care QFTTB testing (QIAGEN, Germantown, MD) at Stony Brook Medicine between October 6, 2020, and March 20, 2024, based on the available institutional data extraction window. Subjects were identified through laboratory records of completed QFTTB testing. All individuals with quantitative TB1 and TB2 results were eligible for inclusion. This study was approved by the Stony Brook Medicine institutional review board (IRB2024-00149) as non-human subjects research. As this was a retrospective study, informed consent requirements were waived. All extracted data were de-identified to ensure subject privacy and confidentiality.

Subjects were excluded if they had incomplete QFTTB data (e.g., missing TB1, TB2, or overall result) or if QFTTB resulted as indeterminate. For those who underwent serial testing, discordance was defined as either conversion (negative-to-positive result) or reversion (positive-to-negative result).

Demographic and clinical characteristics were examined, including age, sex, race/ethnicity, and medical comorbidities such as infection status with Human Immunodeficiency Virus (HIV), cardiovascular disease, autoimmune disease, diabetes, chronic obstructive pulmonary disease (COPD), chronic kidney disease (CKD), and cirrhosis. Medication history was also recorded, including immunosuppressive therapy (e.g., Tumor Necrosis Factor (TNF)-α inhibitors, corticosteroids), cardiovascular medications and pain management drugs. QFTTB test data included dates of testing, quantitative TB1 and TB2 antigen responses, and categorical test result (positive or negative). Quantitative IFN-γ levels were reported in international units per milliliter (IU/mL), the standard output of the QFTTB assay.

Sociodemographic and clinical correlates of IFN-γ responses were first analyzed in the total study population, and subset analysis was carried out in subjects who underwent serial testing stratified by “concordant” and “discordant” results. Descriptive statistics were used to summarize continuous variables (e.g., TB1/TB2 levels) and are reported as means with standard deviations and compared using Welch’s t-tests. Categorical variables (e.g., QFTTB qualitative test result, comorbidities, discordance type) were summarized as frequencies and analyzed using Chi-square or Fisher’s exact tests. Nil, Mitogen, TB1 and TB2 results were correlated with age, sex, race/ethnicity, comorbidities, TB exposure history, TB treatment status, medications and duration between testing using univariate linear regression analyses. Quantitative changes in TB1 and TB2 values were calculated by subtracting baseline from follow-up values. Time intervals between serial tests were calculated in days.

Discordance analysis of the change in TB1 and TB2 values compared converters and reverters using paired t-tests and Wilcoxon signed-rank tests, depending on data distribution. One-way ANOVA was used for comparisons across more than two groups when appropriate. The impact of comorbidities and medication classes on changes in NIL, TB1, TB2, and Mitogen values was assessed using two-way ANOVA, to evaluate both the main effects (discordance group and comorbidity/medication type) and their interaction. The association between discordance type and the presence of specific comorbidities (e.g., HIV, cardiovascular disease, autoimmunity) was evaluated using Chi-square tests. Serial TB1 and TB2 test values within the same donors over time were assessed using linear regression models.

Statistical analysis and data visualization were conducted using RStudio (version 2024.04.2+764; RStudio, PBC, Boston, MA). Statistical significance was assessed using thresholds of *p* < 0.05 and *p* < 0.01, as indicated. No *a priori* power calculation was conducted given the retrospective nature of this study. However, the inclusion of all QFTTB positive cases (n = 436) over a 3.5-year window, along with a matched sample of negative controls, maximized the statistical power for detecting differences in rare outcomes, such as test discordance. Based on the final sample size of 743 subjects, the study had >80% power (α = 0.05) to detect medium effect sizes (Cohen’s *d* ≈ 0.5) in TB1 and TB2 responses between discordance groups. The study period was selected to ensure capture of serial QFTTB testing and representative comorbidities for the subgroup analyses.

## Results

### Study Population

Out of 11,641 QFTTB tests ordered, a total of 436 (3.7%) cases tested positive, and a random age- and sex-matched convenience sample of 307 QFTTB-negative controls was selected from a pool of 10,959 negative QFTTB results. An additional 243 cases did not appropriately respond to the mitogen positive control and TB1 and TB2 testing were not performed (“indeterminate” QFTTB results) and were excluded from the analysis. A total of 743 subjects who underwent QFTTB testing at Stony Brook Medicine between October 6, 2020, and March 20, 2024 were included in the study. IGRA testing was most commonly ordered for employee health screening, including Veterans Home staff (50.8%), followed by renal transplant preoperative evaluation (12.2%), emergency department visits with symptoms (9.1%) or without symptoms (8.8%), obstetric evaluations (3.9%), and the remaining during routine clinical care including HIV, rheumatology, outpatient infectious disease and other advanced specialty clinics. The mean age was 47.0 years (SD ±16.5), with 40.1% identifying as male and 59.1% as female. The racial/ethnic composition included 47.97% White, 8.94% Asian, 20.46% Black/African American, 22.09% Hispanic/LatinX, and 0.54% American Indian or Alaskan Native (**Table 1**). Of the QFTTB+ cases, 16 (3.67%) were diagnosed with microbiologically confirmed active TB infection and the remaining 420 (96.33%) subjects were diagnosed with asymptomatic, presumed LTBI. 203 subjects had serial QFTTB testing, of which 33 (16.2%) demonstrated discordant results. Among all subjects, 29.4% had cardiovascular disease, making it the most prevalent comorbidity in this cohort. Cancer was diagnosed in 12.5% of subjects, 9.2% had COPD, 6.5% were living with HIV and 3.0% had viral hepatitis (Hepatitis B or C viruses) (**Table 1**).

**Table 1.**
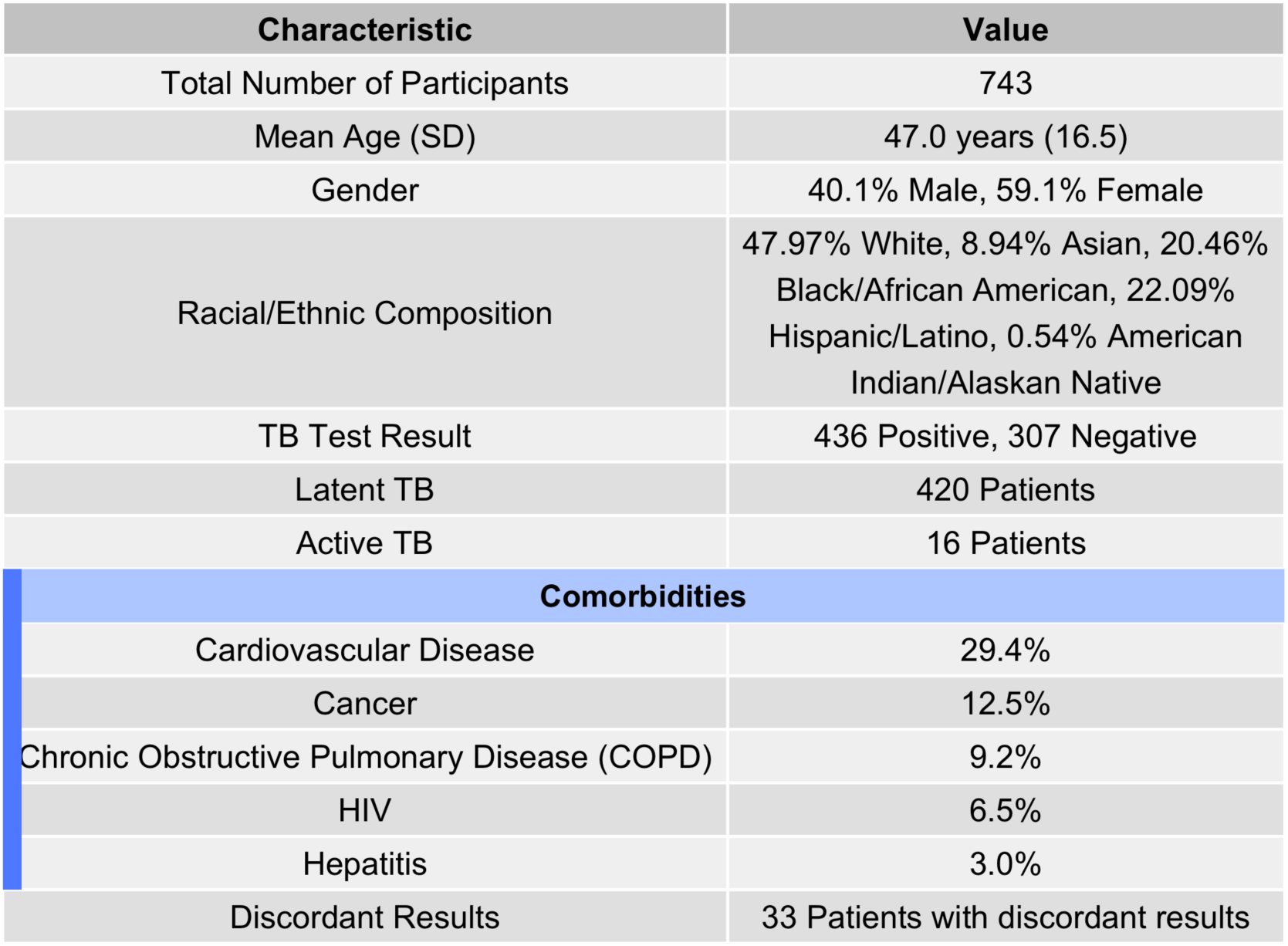
Sociodemographic and Clinical Characteristics of the Study Group. Sampling Method: This dataset represents QFTTB tests conducted at Stony Brook Medicine from **October 6, 2020** to **March 20, 2024**, where all positive cases (n = 436) were included in the analysis and compared to a convenience sample of 307 age- and sex-matched QFTTBnegative controls. All QFTTB testing performed in these subjects during the study period was included in the analysis.

| Characteristic | Value |
| --- | --- |
| Total Number of Participants | 743 |
| Mean Age (SD) | 47.0 years (16.5) |
| Gender | 40.1% Male, 59.1% Female |
| Racial/Ethnic Composition | 47.97% White, 8.94% Asian, 20.46% Black/African American, 22.09% Hispanic/Latino, 0.54% American Indian/Alaskan Native |
| TB Test Result | 436 Positive, 307 Negative |
| Latent TB | 420 Patients |
| Active TB | 16 Patients |
| **Comorbidities** |  |
| Cardiovascular Disease | 29.4% |
| Cancer | 12.5% |
| Chronic Obstructive Pulmonary Disease (COPD) | 9.2% |
| HIV | 6.5% |
| Hepatitis | 3.0% |
| Discordant Results | 33 Patients with discordant results |

Among all study subjects, no significant correlations were observed between TB1-NIL, TB2-NIL, Mitogen-NIL, or NIL values and sociodemographic or clinical variables (**Supplemental Figure 1**).

### Concordant vs. Discordant IGRA Testing

A total of 170 (83.74%) individuals had concordant QFFTB results upon serial testing—89 with persistently negative results and 81 with persistently positive results. Thirty-three subjects (16.26%) had discordant results. The concordant group included 87 females and 83 males; the discordant group included 17 females and 16 males.

There were no statistically significant sociodemographic differences in QFTTB quantitative results between the concordant and discordant serial testing groups (**Figure 1**). The mean age of individuals with concordant results was 45.8 **±**20.0 years, compared to 41.3 **±**16.5 years among discordant subjects (p = 0.1623; 95% CI: –1.87, 10.80) (**Figure 1A**). Sex distribution did not significantly differ between groups (p > 0.05) (**Figure 1B**). The average time between serial QFTTB tests did not significantly differ (p = 0.9423) (**Figure 1C**). No significant differences were observed in TB1 or TB2 changes between concordant and discordant groups (TB1: p = 0.6828; TB2: p = 0.9799) (**Figure 1 D, E**). Scatterplot analysis of IFN-γ concentration changes (TB1 vs. TB2) revealed overlapping patterns between groups (**Figure 1F**).

**Figure 1.**
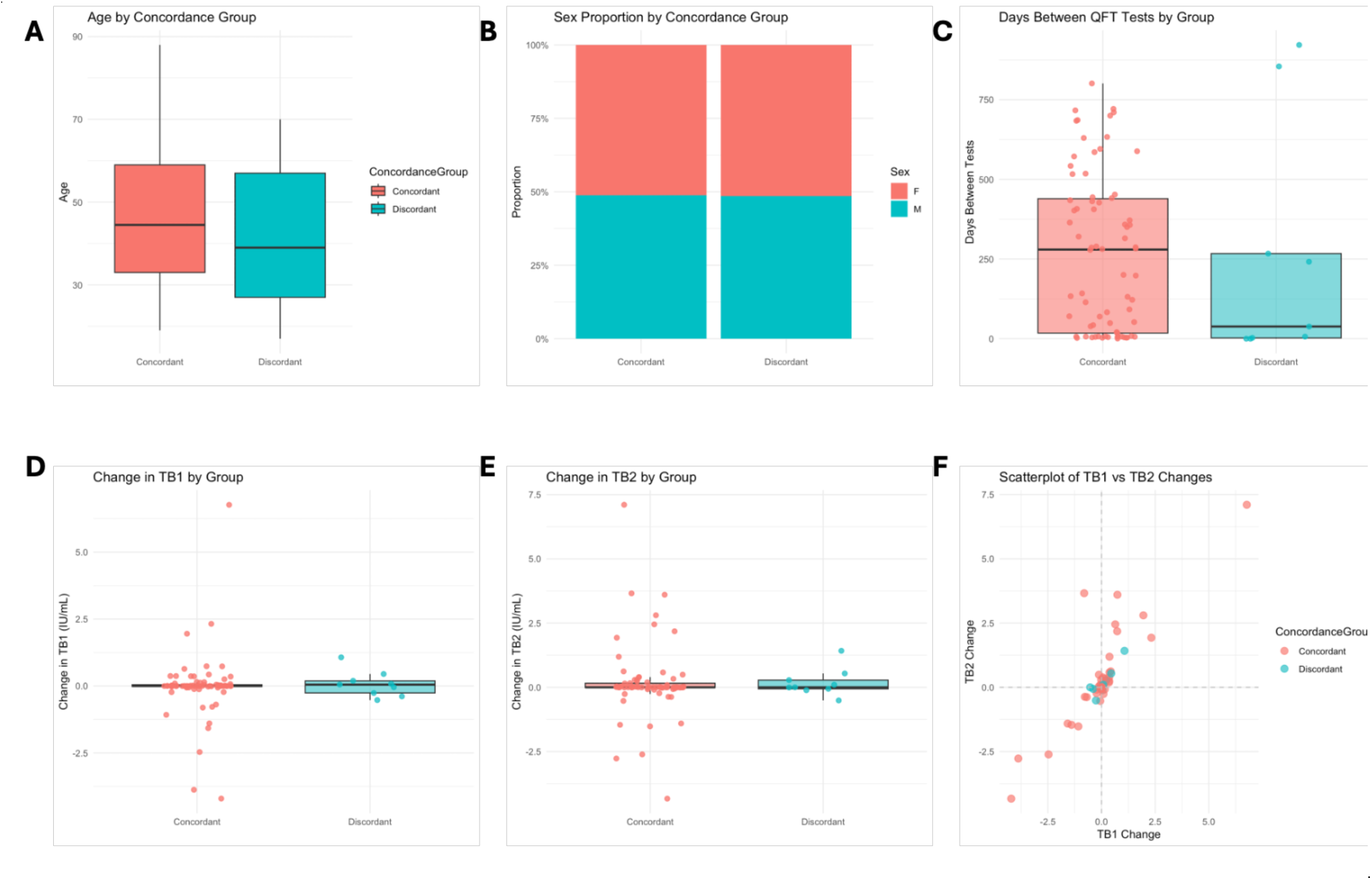
Shared sociodemographic characteristics between serial quantitative QFTTB concordance and discordance groups. **A** Age distribution by concordance group, presented as boxplots. Boxes represent the interquartile range (IQR), horizontal lines denote the median, and whiskers extend to 1.5 × IQR (Welch’s *t*-test *p* = 0.288). **B** Sex distribution by concordance group (concordant vs. discordant), shown as a stacked bar plot. **C** Time in days between serial QFTTB tests by group, shown as boxplots with overlaid individual data points (Welch’s *t*-test *p* = 0.942). **D** Changes in TB1 IFN-γ response by group, displayed as boxplots with jittered individual data points (*p* = 0.683). **E** Changes in TB2 IFN-γ response by group, also shown as boxplots with jittered points (*p* = 0.980). **F** Scatterplot of TB1 vs. TB2 changes illustrates overlapping distributions across groups. Statistical comparisons were performed using Welch’s *t*-tests. No comparisons reached statistical significance.

### Discordant IGRA Testing

Of the 33 study subjects with discordant results, 23 (69.7%) experienced reversion (positive to negative), while 10 (30.3%) experienced conversion (negative to positive). Five (15.2%) of these discordant cases received LTBI treatment with rifampin or isoniazid/rifapentine at the time of testing; 3 in the conversion group and 2 in the reversion group. Average TB1 and TB2 changes in the reversion group were −1.11 ± 0.63 and −1.16 ± 0.42, respectively. In contrast, conversion cases demonstrated average increases of only 0.38 ± 0.17 and 0.37 ± 0.15, respectively.

There was no significant association between sex and conversion measured by Fisher’s exact test (p = 0.4646; odds ratio was 0.5234 (95% CI: 0.0837, 2.9271) (**Figure 2A**). Conversion cases were significantly older than reversions (mean age 51.1 ± 15.0 vs. 37.0 ± 15.6 years; t(17.79) = 2.44, p = 0.025; 95% CI: 1.95–26.16) (**Figure 2B**). Significantly more time elapsed between discordant results for conversions (415.1 days (SD ± 394) vs 91.2 days (SD ±196) (Welch’s test (t(10.927) = 2.5794, p = 0.02574)) (**Figure 2C**).

**Figure 2.**
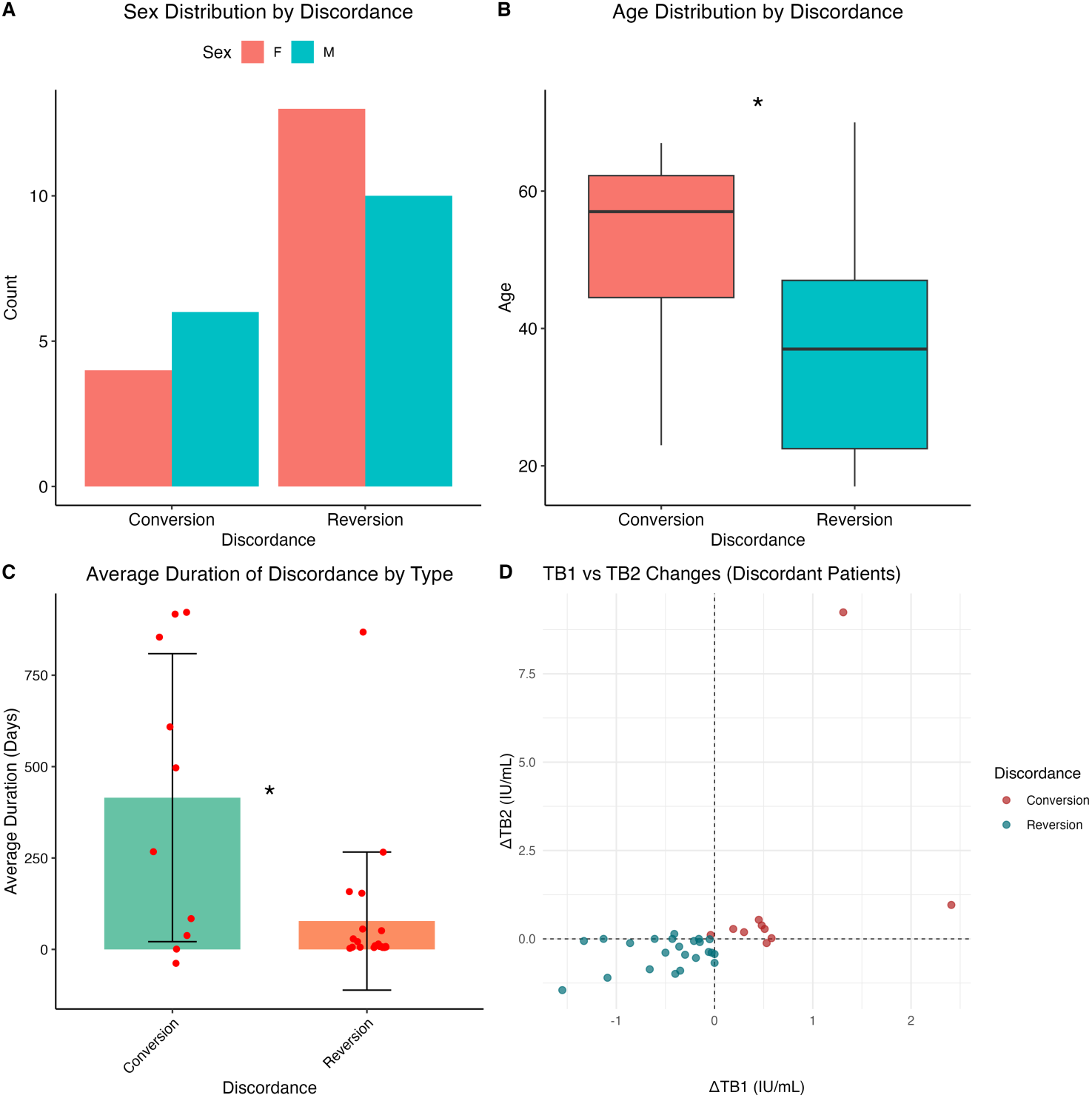
Age and duration between serial testing correlate with QFTTB discordance. **A.** Sex distribution by discordance group (conversion vs. reversion), presented as a bar plot. **B.** Age distribution by discordance group, shown as boxplots. Boxes represent the interquartile range (IQR), horizontal lines denote the median, and whiskers extend to 1.5 × IQR (Welch’s t-test *p =* 0.025). **C.** Mean time in days between serial QFTTB tests by discordance group, shown as a bar plot with standard deviation (SD) error bars (*p* = 0.02574). **D.** Scatterplot displaying changes in TB1 and TB2 IFN-γ responses among individuals with discordant QFTTB results. Statistical comparisons were performed using independent Welch’s t-tests. Asterisks denote levels of statistical significance: *p* < 0.05 (\**), p <* 0.01 *(**)*.

Although converters and reverters are defined by opposing shifts in IFN-γ concentration, the magnitude and distribution of these changes distinguished discordant groups (**Figure 2D**). Converters exhibited a wider range of increases in TB1 (median +0.495 IU/mL, range –0.04 to +2.41) and TB2 (median +0.28 IU/mL, range –0.12 to +9.24), with statistical outliers identified at 2.41 IU/mL (TB1) and 9.24 IU/mL (TB2) based on a z-score threshold > 2.5. In contrast, reverters demonstrated more uniform decreases in TB1 (median –0.36 IU/mL, range –1.55 to 0.00) and TB2 (median –0.37 IU/mL, range –1.45 to +0.14), with over half demonstrating TB2 decreases greater than 0.6 IU/mL.

### Impact of Clinical Factors on Discordant Results

We next examined the relationship between comorbidity and changes in NIL, TB1, TB2, and mitogen responses by discordance group (**Figure 3A–D**). Comorbidity was significantly associated with changes in NIL (**Figure 3A**), TB1 (**Figure 3B**) and most notably TB2 (F(3, 8) = 275.64, p = 1.75 × 10⁻⁷) (**Figure 3C**), but not discordance (F(3, 8) = 4.90, p = 0.058).

**Figure 3.**
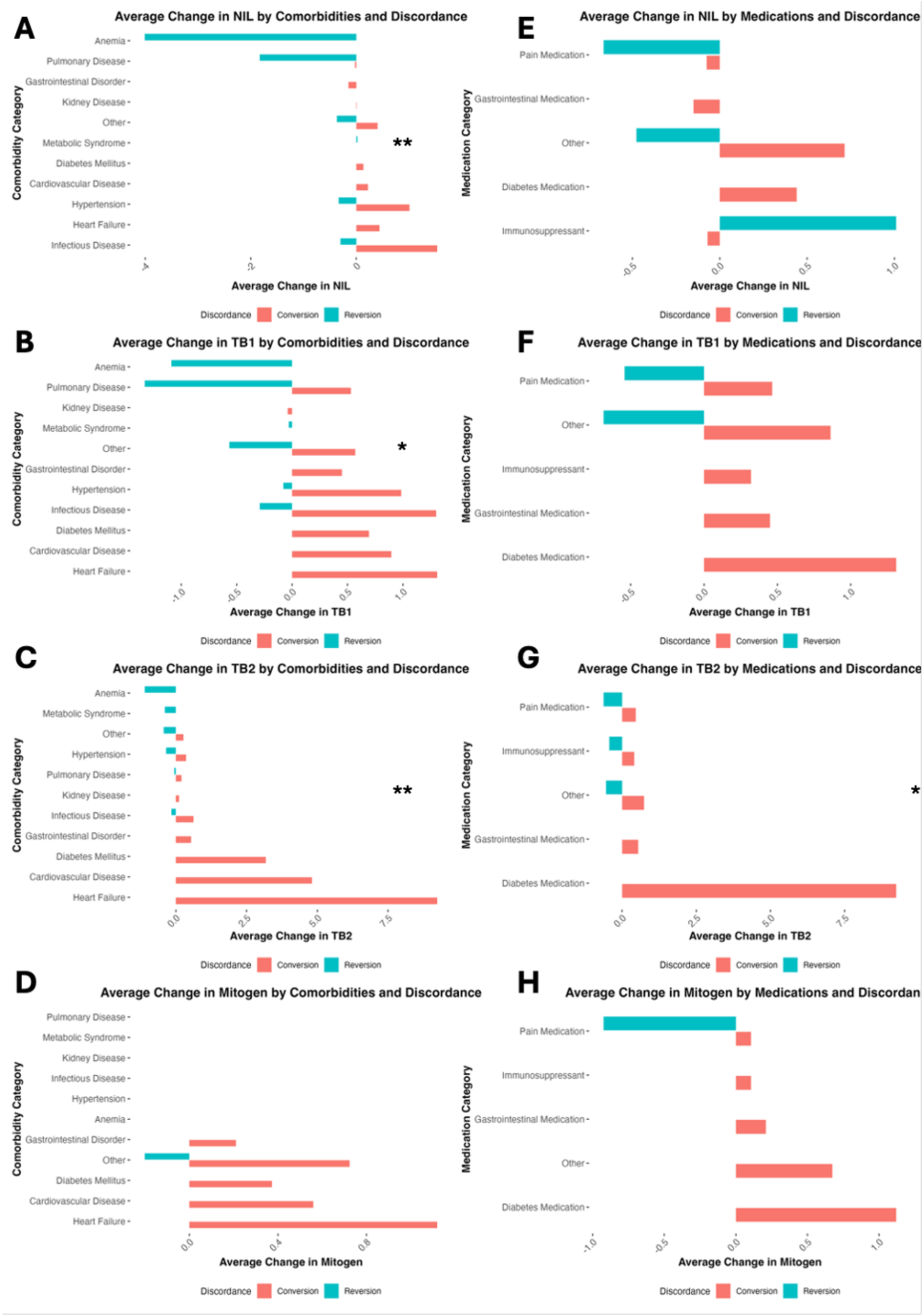
Clinical comorbidities and medications correlate with change in quantitative QFTTB result. Bar plots showing the average change in NIL (**A**), TB1 (**B**), TB2 (**C**) and Mitogen (**D**) stratified by comorbidity and discordance group (conversion vs. reversion). Bar plots showing the average change in NIL (**E**), TB1 (**F**), TB2 (**G**), and Mitogen (**H**) stratified by medication category and discordance group. Statistical comparisons were performed using a two-way ANOVA. Asterisks denote levels of statistical significance: *p* < 0.05 (\**), p <* 0.01 *(**).*

Comorbidities were not significantly associated with mitogen responses (F(3, 8) = 0.025, p = 0.994) (**Figure 3D**), indicating preserved T cell function across groups.

Average positive changes in IFN-γ concentration in the NIL condition were highest in individuals with hypertension or infectious diseases, while negative changes correlated with mild anemia or pulmonary disease (**Figure 3A**). For TB1, the largest positive changes were observed in individuals with diabetes mellitus or cardiovascular disease, while the largest negative changes were seen in those with anemia or pulmonary disease (**Figure 3B**). TB2 responses showed the greatest positive changes in conversion subjects with cardiovascular disease or diabetes mellitus, and the largest negative changes in reverters with pulmonary disease (**Figure 3C**). Mitogen responses were relatively similar across comorbidity groups, with no pronounced directional pattern (**Figure 3D**). We also assessed the effects of medication on QFTTB results (**Figure 3E–H)**. No significant associations were observed between medication use and changes in NIL (**Figure 3E**), TB1 (**Figure 3F**), or mitogen responses (**Figure 3H**). However, TB2 positive changes were significantly associated with medication class (F(1, 3) = 26.71, p = 0.014), with highest positive changes among converters on diabetes medications, such as metformin, and gastrointestinal medications (**Figure 3G**).

We observed differences in comorbidity burden between the discordant groups. Among the 23 reversion cases, only 5 (22%) had a serious chronic condition (e.g., HIV, End stage renal disease), while 18 (78%) had no comorbidities or mild disease (e.g., anemia, dyslipidemia, obesity or anxiety). In contrast, 8 of 10 converters (80%) had at least one significant chronic disease (e.g., HIV, diabetes, or cardiovascular disease).

## Discussion

Our single-center study provides important insights into the performance of QFTTB testing in a low-incidence setting for TB transmission. We found that 16.2% of subjects undergoing serial QFTTB screening demonstrated discordant results. Five percent of subjects with initial positive testing underwent confirmatory testing and demonstrated discordant negative results. Our analyses revealed that reverters were significantly younger and underwent retesting over shorter intervals. In contrast, converters were older and had more chronic comorbidities and medication use. We conclude that discordant QFTTB results are common and confound accurate assessment of TB risk. While three-month weekly isoniazid-rifapentine or four-month daily rifampin regimens that are currently first-line therapy for LTBI^22^ are generally well-tolerated, they may introduce significant side effects including gastrointestinal intolerance, hepatitis, drug-induced hypersensitivity and gut dysbiosis^23,24^. As such, interpretation of QFTTB testing results may warrant referral to an Infectious Diseases specialist to stratify risk and evaluate utility of LTBI treatment.

A notable finding in our study was the significant age difference between individuals in the conversion and reversion groups with a mean age 51.1 vs. 37.0 years at most recent testing (*p* = 0.0253). This age difference does not appear to be driven by the duration between serial testing (415.1 days (SD ± 394) vs 91.2 days), which was on average less than 2 years of follow-up per case. The majority of reverters had no significant clinical comorbidities. Younger individuals underwent repeat QFTTB testing over shorter intervals, often in occupational or routine screening contexts, where 5.28% of initially positive results reverted to negative upon retesting. This shorter testing interval among reverters likely represents reflex retesting triggered by low pre-test probability or occupational screening guidelines, where confirmation is recommended. Subjects undergoing TB testing despite low pre-test probability of infection are at increased risk for false-positive tests^25^. We believe this is the most likely explanation for the inverse association between reversion and patient age. Our results underscore the importance of existing guidelines offered by the Centers for Disease Control and Prevention to repeat QFTTB testing if the probability of *Mtb* exposure and LTBI are low prior to considering treatment^26^.

In contrast, converters had longer intervals between testing that was significantly associated with chronic comorbidities and medication use, namely metformin that correlated with highest change in TB2 values. This suggests that chronic illness and medications used may influence QFTTB responses. Additionally, chronic diseases may be associated with persistent low-grade inflammation, potentially augmenting immune responses and subsequent QFTTB immunoreactivity^27,28^. Older subjects also experience increased cumulative risk of *Mtb* exposure over time, including frequent contact with the healthcare system or foreign travel, that represent potential confounding variables not captured in our study. As of 2024, Suffolk County, New York reported an active TB case rate of 3.4 per 100,000 annually (5.4 per 100,000 in New York State), making local transmission unlikely^21^. Our results underscore the importance of integrating clinical history and epidemiologic risk when ordering or interpreting QFTTB testing in low-risk settings.

We acknowledge that additional biological variables that may modulate QFTTB reactivity were not measured in this study. In rare cases, false positives may result from immunologic cross-reactivity with non-tuberculous mycobacterial antigens derived from *M. kansasii* or *M. marinum*^32^. Diurnal fluctuation, transient inflammation, recent infections, or vaccinations may also modulate immune responses and QFTTB reliability^5,33^. Operational variables such as sample handling procedures and quality control are also potential contributors to the observed discordance, which could not be measured in this study. In the US, QFTTB testing is required to be carried out in Clinical Laboratory Improvement Amendments (CLIA’88) compliant laboratories that adhere to regulations from bodies such as the College of American Pathology.

This requires adherence to operational temperature ranges and standard operating procedures (SOP). Deviations from SOPs are flagged in the EMR and results withheld if criteria are not met. Thus, the results reported pass rigorous quality assurance protocols prior to release. However, operational and pre-analytical variables including delays in sample transport to lab and temperature instability during transport and storage may impact test reproducibility^4,19,33,34^. As samples must be processed within 12 hours of collection^35^, variability in time to process may contribute to assay fluctuation^36,37^. There are also potential operator-dependent variables in the application of the test itself including initiation of the procedure, improper tube mixing, and variable incubation period prior to quantitation^33,37^. Discordance may be influenced by any of these operational variables that could not be captured in the study.

We also recognize the limitation of this being a single-center retrospective analysis of QFTTB testing in a region with low TB incidence that did not evaluate alternative IGRA platforms. Thus, we expect that our results will be most relevant to other low-prevalence clinical settings in which individuals undergo serial QFTTB testing. However, we also believe that our findings may apply to high incidence settings where serial QFTTB is being used as surrogate endpoint for incident *Mtb* infection in TB vaccine clinical trials^10,29^. ^10,30,31^. Discordant serial testing that does not accurately reflect *Mtb*-specific responses can confound both clinical decision-making and interpretation of clinical trial results.

In sum, our results demonstrate significant variability in QFTTB results during serial testing that should raise caution when interpreting positive results and considering LTBI therapy initiation. Coordination with local clinical laboratories, confirmatory testing and Infectious Diseases specialist consultation may be helpful in assessing these cases. Ongoing work seeks to define the immunologic mechanisms underlying hypothesized non-specific reactions to *Mtb* peptide pools that may be driving QFTTB discordance. This includes examining the relative contribution of alternative IFN-γ-secreting immune subsets such as natural killer cells and innate-like T cells relative to conventional, *Mtb* peptide-specific CD4^+^ and CD8^+^ T cells that the assay is designed to target.^38,39^.

## Data Availability

All data produced in the present study are available upon reasonable request to the authors.

## Acknowledgments

We are thankful for the support of the Stony Brook Foundation, the Department of Medicine of the Renaissance School of Medicine and the Office of the Vice President of Research of Stony Brook University.

## Author contributions

The following authors contributed to the manuscript in various ways. Each author’s contribution is listed below.

Conceptualization: GKS, AT, LAM, CKV

Methodology: GKS, AT, LAM, CKV

Investigation: GKS and CKV

Visualization: GKS and CKV

Funding acquisition: CKV

Writing – original draft: GKS and CKV

Writing – review & editing: all authors

---

**Supplemental Figure 1.**
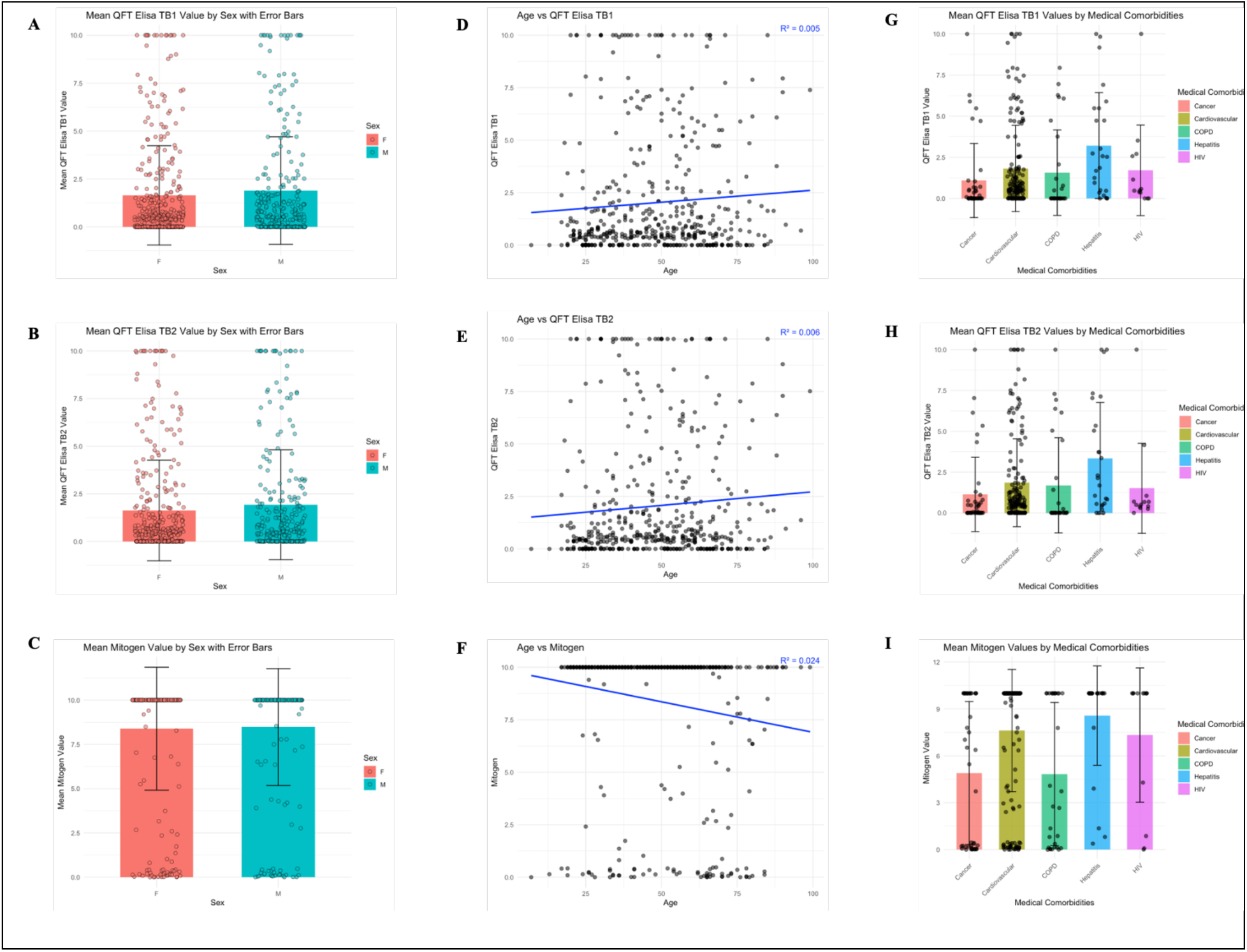
Distribution of QFTTB IFN-γ responses by sex, age, and medical comorbidities. A–C: TB1 IFN-γ values stratified by sex (A), plotted against age with linear regression (B), and shown across comorbidity categories (C). TB2 IFN-γ values stratified by sex (D), plotted against age with linear regression (E), and shown across comorbidity categories (F). Mitogen IFN-γ values stratified by sex (G), plotted against age with linear regression (H), and shown across comorbidity categories (I). Each panel displays mean IFN-γ concentrations (IU/mL) as measured by QFTTB component assays. Statistical comparisons were performed using linear regression or ANOVA as appropriate.

